# The impact of Illinois’ comprehensive handheld phone ban on talking on handheld and handsfree cellphones while driving

**DOI:** 10.1101/2022.04.08.22273533

**Authors:** Marco H. Benedetti, Bo Lu, Neale Kinnear, Li Li, M. Kit Delgado, Motao Zhu

## Abstract

**Introduction:** Distracted driving has been linked to multiple driving decrements and is responsible for thousands of motor vehicle fatalities annually. Most US states have enacted restrictions on cellphone use while driving, the strictest of which prohibit any manual operation of a cellphone while driving. Illinois enacted such a law in 2014. To better understand how this law affected cellphone behaviors while driving, we estimated associations between Illinois’ handheld phone ban and self-reported talking on handheld, handsfree, and any cellphone (handheld or handsfree) while driving.

**Methods:** We leveraged data from annual administrations of the Traffic Safety Culture Index from 2012-2017 in Illinois and a set of control states. We cast the data into a difference-in-differences (DID) modeling framework, which compared Illinois to control states in terms of pre-to post-intervention changes in the proportion of drivers who self-reported the three outcomes. We fit separate models for each outcome, and fit additional models to the subset of drivers who talk on cellphones while driving.

**Results:** In Illinois, the pre-to post-intervention decrease in the drivers’ probability of self-reporting talking on a handheld phone was significantly more extreme than that of drivers in control states (DID estimate −0.22; 95% CI −0.31, −0.13). Among drivers who talk on cellphones while driving, those in Illinois exhibited a more extreme increase in the probability of talking on a handsfree phone while driving than those control states (DID estimate 0.13; 95% CI 0.03, 0.23).

**Conclusions:** Our results suggest that Illinois’ handheld phone ban reduced talking on handheld phones while driving and corroborated the hypothesis that the ban promoted harm-reduction via substitution from handheld to handsfree phones among drivers who talk on the phone while driving.

**Practical Applications:** Our findings should encourage other states to enact comprehensive handheld phone bans to improve traffic safety.

## 1: INTRODUCTION

Distracted driving is an ongoing public health issue in the United States.^1^ The National Highway Traffic Safety Administration estimated that 3,142 people were killed in motor vehicle crashes (MVCs) involving distracted drivers in 2019, with another approximately 424,000 injured.^2^ A common form of distracted driving, and the focus of the present study, is talking on a cellphone while driving. Taking on a handheld cellphone while driving is associated with several decrements to driving performance, including delayed reaction time, increased lateral lane positioning, and failure to maintain proper speed and distance.^3,4^ These can occur due to both cognitive distractions and concurrent manual tasks, such as dialing or reaching for the cellphone.^5^

To curtail distracted driving in the United States, all but two states have enacted laws that restrict all drivers from using a handheld cellphone while driving in some fashion.^6^ Twenty-four states ban talking on a handheld cellphone while driving, with the most comprehensive of these laws prohibiting all manual operation of a handheld cellphone while driving. On January 1, 2014, Illinois placed into effect such a law. Specifically, Illinois Compiled Statute 12-610.2 states, “A person may not operate a motor vehicle on a roadway while using an electronic communication device, including using an electronic communication device to watch or stream video.” An electronic communication device is an “electronic device, including, but not limited to, a hand-held wireless telephone, hand-held personal digital assistant, or a portable or mobile computer, but does not include a global positioning system or navigation system or a device that is physically or electronically integrated into the motor vehicle.” Several specific exceptions are present in the law, including the use of an electronic communication device for emergencies (ICS 12-610.2 Section (d)-2) and handsfree devices (ICS 12-610.2 Section (d)-3).^7^

Previous studies have shown that handheld phone bans reduce handheld phone use while driving^8-18^ and that they are associated with reductions in traffic fatalities,^19-22^ although the latter association has seen mixed evidence.^23^ However, it is less clear how handheld phone bans associate with handsfree phone use while driving. With US states continuing to change handheld phone laws, and the availability of handsfree phone technology increasing concurrently, it is pertinent to study how handheld phone bans affect driver behaviors with respect to both existing and evolving communication technology. Carpenter and Nguyen proposed the “substitution hypothesis” about handheld and handsfree phone use based on a quasi-experimental analysis of a handheld phone ban in Ontario, Canada.^10^ They found that the ban was associated not only with less handheld phone use, but also more handsfree phone use, suggesting that the ban deterred handheld phone use and lead drivers to substitute a handheld phone for a handsfree one. This hypothesis was corroborated by Benedetti et al. (2022), who employed a cross-sectional design and regression analysis to estimate the association between handheld phone bans and self-reported talking on handheld, handsfree, and any cellphone while driving in the United States.^24^ While Benedetti et al. benefitted from a large and nationally representative national data set, their cross-sectional study design was not as well-suited to identify causal effects as the quasi-experimental study of Carpenter and Nguyen.

To our knowledge, there is currently no US equivalent to Carpenter and Nguyen’s quasi-experimental study of handheld and handsfree phone use in Canada. This study aims to fill that research gap and complement the cross-sectional study of Benedetti et al. (2022) Specifically, we cast data from annual implementations of the Traffic Safety Culture Index survey into a difference-in-differences framework to estimate the effects of Illinois’ comprehensive handheld phone ban on self-reported talking on (i) handheld phones; (ii) handsfree phones; and (iii) any cellphone, handheld or handsfree, while driving. Based on the findings of Carpenter and Nguyen and Benedetti et al., and rooted in the substitution hypothesis, we expect that the ban will be associated with less talking on handheld phones and more talking on handsfree phones, as well as less talking on any cellphone, handheld or handsfree.

## 2: METHODS

### 2.1: Study Data

The Traffic Safety Culture Index (TSCI) is an annual survey administered by the AAA Foundation for Traffic Safety.^25^ Administered to approximately 3,500 Americans annually, it collects information on various self-reported behaviors and attitudes about traffic safety, including distracted driving. Due to the availability of survey items relevant to our research questions, we used surveys from years 2012 through 2017. We included drivers aged 21 and older to avoid potential confounding due to young-driver cellphone bans. In planning a quasi-experimental analysis of handheld phone bans, we needed to identify states that enacted handheld phone bans during this time frame. From 2012-2017, four states enacted handheld phone bans: Illinois, Vermont, Hawaii, and New Hampshire. However, only Illinois had enough observations (233 pre-law, and 497 post-law) to be analyzed on its own. All data were anonymized and therefore exempt from IRB review at Nationwide Children’s Hospital.

### 2.2: Inclusion Criteria and Causal Assumptions

We wished to compare the pre-to-post-intervention trends in talking on handheld, handsfree, and any cellphone (handheld or handsfree) from Illinois to those of a set of control states. Prior to enacting their comprehensive ban on handheld phone use while driving, Illinois prohibited manual typing or reading text on a handheld device. Candidate control states needed to have a similar law in place for the duration of the study. Twenty-four states met this criterion: Twenty-four states met this criterion: Arkansas, Colorado, Georgia, Idaho, Iowa, Indiana, Kansas, Kentucky, Louisiana, Maine, Maryland, Massachusetts, Minnesota, Michigan, Nebraska, North Carolina, North Dakota, Pennsylvania, Rhode Island, Tennessee, Utah, Virginia, Wisconsin, and Wyoming,.

To facilitate a valid comparison between Illinois and controls states, we applied additional inclusion criteria. Our goal was to select control states that reasonably represent the counterfactual scenario in which Illinois had not enacted their comprehensive handheld phone ban, which is critical in our ability to attribute the estimated effects to Illinois’ handheld phone ban. Multiple causal assumptions are usually needed for this to be the case. The first one is stable unit treatment value assumption (SUTVA). In the context of our study, it suggests that there were no spillover effects of Illinois handheld phone ban. The second assumption is the ignorability assumption, which has a key component of “unconfoundedness.” It asserts that, after conditioning on a complete set of confounders, subjects in both Illinois and control states would exhibit similar outcomes if they were subject to the same handheld phone ban policy.

Additional details can be found in, for example, Rosenbaum, 2010.^26^ These conditions in tandem are closely related to the parallel trends assumption, which is often invoked for quasi-experimental studies such as this one. The parallel trends assumption says that in the absence of an intervention, the difference between Illinois and the control states would have remained constant over time (after controlling for confounding factors). It cannot be proven formally, however we attempted to make a strong of a case as possible for parallel trends via a two-pronged approach. First, our model adjusts for several demographic and economic factors derived from TSCI survey items (see **Sections 2.3** and **2.4**). Second, acknowledging that large-scale regional differences in the United States may contribute to additional unmeasured confounding that we could not account for in a regression model, we constructed a control group consisting only of states that either (i) border Illinois, or (ii) belong to the Midwest Census Region. While this approach had potential to induce spillover effects, we assessed that the potential for such effects was small (see **Section 4.1**), therefore we prioritized controlling for unmeasured confounding when selecting control states. Applying the inclusion criteria, our control states were Indiana, Iowa, Kentucky, Minnesota, Michigan, and Wisconsin.

### 2.3: Study Variables

We analyzed three outcome variables: (i) self-reported talking on a handheld phone while driving; (ii) self-reported talking on a handsfree phone while driving; and (iii) self-reported talking on any cellphone (either handheld or handsfree) while driving in the past month. For the former two outcomes, we presented results for all drivers and for the subset of drivers who talk on any type of cellphone while driving. All variables were derived from questions in the TSCI surveys. Likert-scale responses were converted to binary indicators of an affirmative response, i.e., a value of 1 indicates that the respondent talked on a handheld/handsfree/any cellphone at least once while driving in the last month.

We leveraged two binary exposure variables to estimate the association between Illinois’ handheld phone ban and the three outcomes. The first was an indicator of whether the subject took their survey before or after the passing of Illinois handheld phone ban on 1/1/2014. The second was an indicator of a subject’s state of residence, either in Illinois or not. Although we sought to select control states that would alleviate potential, especially unmeasured, confounding, there were nonetheless demographic differences between Illinois and the control states. Therefore, our analysis incorporated the following demographic/economic variables from TSCI survey items: gender, age, income, education, race/ethnicity, metropolitan status, and household size. An assessment of these covariates’ balance between Illinois and control states before and after the handheld phone ban affirms that this adjustment was appropriate (**Table S1** in the Supplemental Materials). Furthermore, the demographic covariates comprised all factors used in sample selection and weighting, thereby improving our ability to infer about our target population.^27,28^

### 2.4: Statistical Analysis

We began with a descriptive analysis consisting of cross-tabulations of relevant study variables. First, we generated percentages of the outcomes for both Illinois and control states, both before and after Illinois’ handheld phone ban was enacted. Next, to assess covariate balance between the control and treatment group, we provided cross-tabulations of demographic variables by treatment group before and after Illinois’ law was enacted.

To estimate the differences between Illinois and controls in terms of pre-to-post-intervention change in the outcomes, we cast individual-level TSCI data into a difference-in-differences (DID) modeling framework,^29^ the linear component of which is stated below:

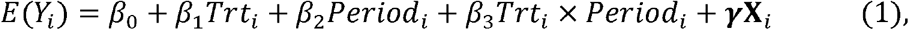

where *Y*_*i*_ is one of our three binary response variables for subject i; *E*(·) denotes the expectation; *Trt*_*i*_ is equal to 1 if subject i resides in Illinois, and 0 if they live in a control state. *Period*_*i*_ is an indicator variable denoting the time period in which subject *i* took the survey, taking on a value of 0 if subject *i* took the survey before 1/1/2014, and a value of 1 if they took the survey after 1/1/2014. *X* is a matrix of additional demographic covariates listed in Section 2.2.

The critical parameter in equation (1) is *β*_3_, the so-called difference-in-difference parameter. It estimates Illinois’ observed divergence from the control states in terms of their pre-to-post-intervention changes in the outcomes. Under the assumption that the control states represent the counterfactual in which Illinois had not changed their policy, *β*_3_ estimates the causal effect of Illinois’ comprehensive handheld phone ban. Absent this assumption, *β*_3_ is nonetheless a robust estimate of the association between handheld phone bans and the three outcomes.

We modeled the data using the GLM procedure in SAS Ver. 9.4. The procedure employs general estimating equations, which allowed us to ensure that the standard error estimates were robust to residual correlation between subjects in the same state that was not captured through the linear terms in equation (1). Although the data were binary, we modeled their expectation with an identity link function because neither the fitted values from the model nor their corresponding 95% confidence limits were close to falling outside the data’s support from zero to one. This approach facilitated greater ease in interpreting the interaction term on the probability scale.^30^

We are aware of several recent methodological developments in policy evalutaion,^31-34^ many of which build upon the seminal synthetic control method of Abadie et al. (2010).^35^ These methods could have been employed in this setting to create a more suitable control group and alleviate potential bias that has been observed for traditional DID in certain settings. However, we assessed that the regression model based around the traditional DID method was the best approach in our setting because (i) we had few control states and pre-intervention time points available, making the synthetic control method and its extensions untenable; (ii) we augmented our data to only have two time points (before and after the handheld phone ban).^36^ We therefore did not need to account for temporal correlation in the data, which can improve DID inference for longitudinal data.^33,37^

## 3: RESULTS

**Table 1** presents the pre- and post-intervention percentages who self-reported talking on handheld, handsfree, and any cellphone while driving in Illinois and control states. For greater clarity, these statistics are also plotted in **Figure 1**. Temporal trends in each of the outcomes are apparent and often differ between Illinois and control states. Both Illinois and the control states exhibited reductions in self-reported talking on handheld phones while driving during the study period (**Table 1**; **Figure 1 (a)** and **(b)**). However, the reduction in Illinois was noticeably more extreme than in the control states. This was true among all drivers and among the subset of drivers who talk on the phone while driving. The trends in self-reported talking on a handsfree phone while driving among all drivers were similar between Illinois and control states (**Figure 1 (c)**); both increased over the study period. However, among drivers who talked on the phone while driving, Illinois exhibited a more extreme increase in talking on handsfree phones than the control states (**Figure 1 (d)**). Talking on any cellphone (handheld or handsfree) while driving decreased in both Illinois and the control group, however the decrease in Illinois appeared to be more pronounced (**Figure 1 (e)**).

**Table 1:**
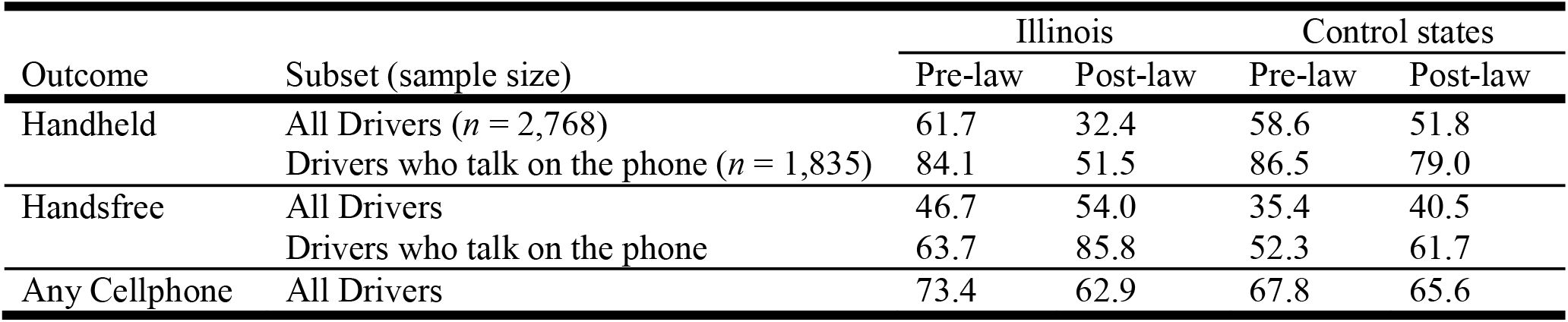
Percentage who self-reported talking on handheld, handsfree, and any cellphone (handheld of handsfree) while driving by law period and time period. “Pre-law” refers to drivers who took the TSCI survey before 1/1/2014, and “Post-law” refers to drivers who took the survey after 1/1/2014. Control states included Indiana, Iowa, Kentucky, Minnesota, Michigan, Kentucky, and Wisconsin.

| Outcome | Subset (sample size) | Illinois |  | Control states |  |
| --- | --- | --- | --- | --- | --- |
|  |  | Pre-law | Post-law | Pre-law | Post-law |
| Handheld | All Drivers ($n = 2,768$) | 61.7 | 32.4 | 58.6 | 51.8 |
| | Drivers who talk on the phone ($n = 1,835$) | 84.1 | 51.5 | 86.5 | 79.0 |
| Handsfree | All Drivers | 46.7 | 54.0 | 35.4 | 40.5 |
|  | Drivers who talk on the phone | 63.7 | 85.8 | 52.3 | 61.7 |
| Any Cellphone | All Drivers | 73.4 | 62.9 | 67.8 | 65.6 |

**Figure 1:**
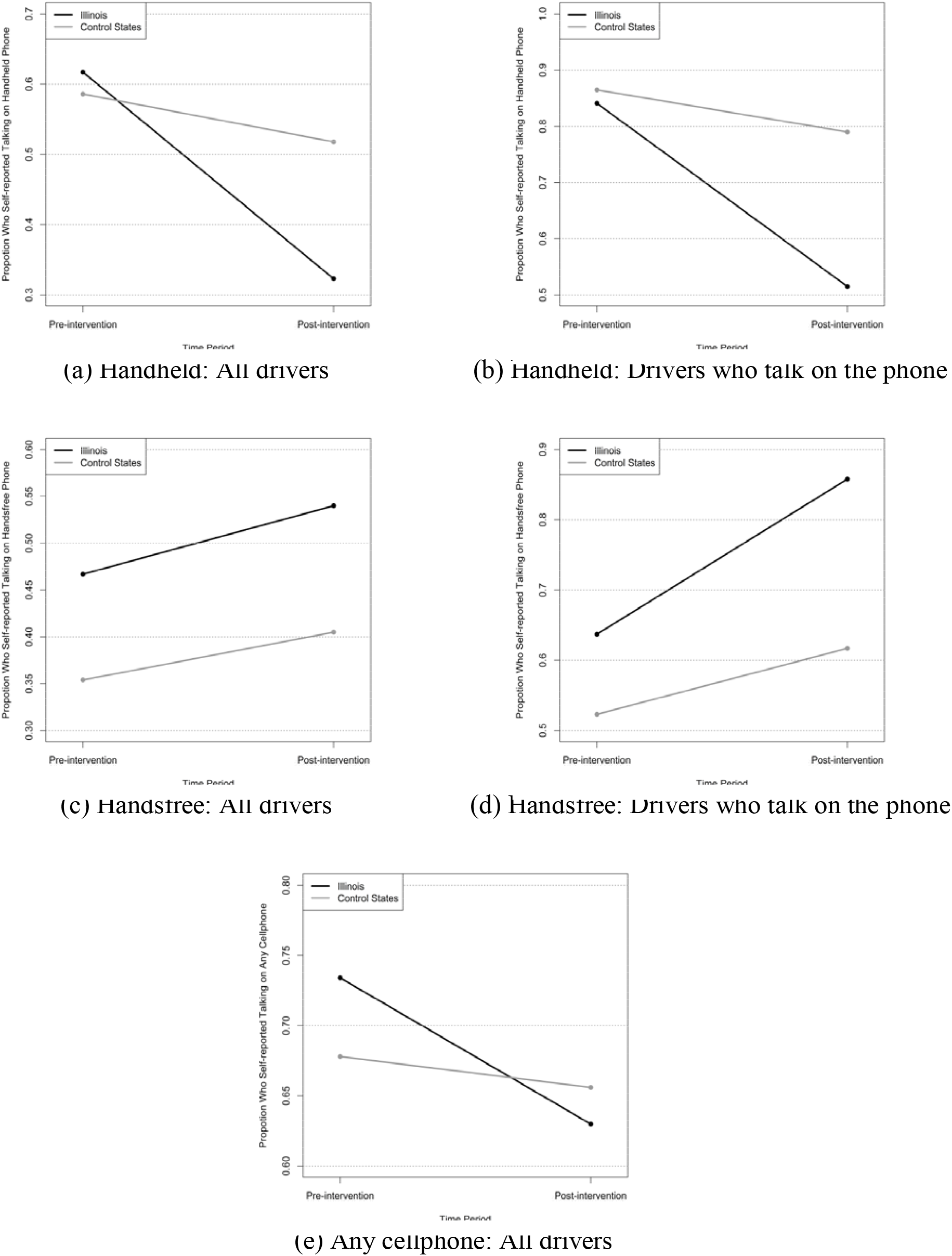
Proportion who self-reported talking on handsfree, handheld, and any cellphone while driving in Illinois before and after they implemented a handheld phone ban.

**Table 2** presents results from our difference-in-differences model. Specifically, we provide our estimates and corresponding 95% confidence intervals (CI) for *β*_3_ in model (1), which measures the difference between Illinois and the control states in terms of their pre-to-post-intervention changes in the outcomes. In Illinois, the pre-to post-intervention decrease in the probability of drivers self-reporting talking on a handheld phone was significantly more extreme than that of drivers in control states (DID estimate −0.22; 95% CI −0.31, −0.13; *p*-value < 0.001). A similar but more extreme difference was observed among the subset of drivers who talk on the phone (DID estimate −0.27; 95% CI −0.36, −0.17; *p*-value < 0.001). Among all drivers, we found no evidence of a differential increase in talking on handsfree phones between Illinois and control states (DID estimate 0.03; 95% CI −0.06, 0.12; *p*-value = 0.525). However, among drivers who talk on the phone while driving, those in Illinois exhibited a more extreme increase in the probability of talking on a handsfree phone while driving than those in control states (DID estimate 0.13; 95% CI 0.03, 0.23; *p*-value = 0.013). The pre-to-post-intervention decrease in the probability that a driver talks on any phone (handheld or handsfree) while driving in Illinois may have been more extreme than that of the control states, however this estimate was not statistically significant at the common significance level of 0.05 (DID estimate −0.07; 95% CI −0.15, 0.05; *p*-value = 0.100).

**Table 2:** Results from fitting the model in equation (1). The column “DID Estimate” corresponds to *β*_3_, which estimates Illinois divergence from control states in terms of pre-to-post-intervention changes in the outcomes. Estimates adjust for gender, age, income, education, race/ethnicity, metropolitan status, and household size.

| Outcome | Subset | DID Estimate<br>(95% CI) | *p*-value |
| --- | --- | --- | --- |
| Handheld | All Drivers | **-0.22 (-0.31, -0.13)** | **<0.001** |
|  | Drivers who talk on the phone | **-0.27 (-0.36, -0.17)** | **<0.001** |
| Handsfree | All Drivers | 0.03 (-0.06, 0.12) | 0.525 |
|  | Drivers who talk on the phone | **0.13 (0.03, 0.23)** | **0.013** |
| Any cellphone | All Drivers | -0.07 (-0.15, 0.01) | 0.100 |

## 4: DISCUSSION

Our study estimated the effects of Illinois’ comprehensive handheld phone ban on self-reported talking on handheld, handsfree, and any cellphone (either handheld or handsfree) while driving. We leveraged a quasi-experimental design and difference-in-differences modeling framework, providing a US-based equivalent to the study of Carpenter and Nguyen (2015) in Canada,^10^ and complementing the cross-sectional study of Benedetti et al. (2022),^24^ which also used TSCI data. To our knowledge, it is the first quasi-experimental study of US data to estimate the effects of handheld phone bans on both handheld and handsfree driving behaviors.

We found strong evidence that Illinois’ handheld phone ban was associated with a reduction in talking on handheld phones while driving, and that this reduction was more extreme than the one observed in control states. This finding was consistent with several other studies supporting the efficacy of handheld phone bans to reduce handheld phone use while driving.^9-18,24^ When considered in the context of evidence that (i) handheld phone use is associated with multiple decrements in driving performance;^3-5^ and (ii) comprehensive handheld phone bans like the one in Illinois are associated reductions in MVC fatalities;^22^ these findings underscore the importance of handheld phone bans improve traffic safety.

Based on previous studies and the substitution hypothesis, we expected that Illinois’ drivers would exhibit a more extreme in talking on handsfree phones while driving increase than drivers in control states. We found practically no evidence of such an association among all drivers. However, among drivers who talk on the phone while driving, Illinois exhibited a significantly more extreme increase in the probability that those drivers will do so on a handsfree phone compared to control states. These results in tandem only partially corroborate the substitution hypothesis, however they suggest that drivers who engage in the potentially risky behavior of talking on cellphones while driving are more likely to do so with a handsfree phone when a handheld phone ban is in place. The idea that handheld phone bans lead drivers to switch from handheld phones to handsfree ones has been interpreted in multiple ways. For example, Carpenter and Nguyen described substitution as an unintended effect of handheld bans,^10^ whereas the authors of the present study previously described substitution as a form of harm reduction (we note that these viewpoints are not necessarily mutually exclusive).^24^ Although simulator studies have identified driving decrements associated with talking on a handsfree phone,^38-41^ naturalistic driving studies have shown that talking on handheld phones and associated manual/visual distractions are more harmful than talking on a handsfree phone.^5,42-44^ While not talking on the phone while driving is the safest behavior, substitution from a handheld to a handsfree phone can be considered as a form of harm reduction from a public health perspective.

Benedetti et al. found that handheld phone bans like the one in Illinois were associated with significantly lower talking on any cellphone (handheld or handsfree), suggesting that, despite potential substitution, handheld phone bans were associated with a net reduction in talking on the phone while driving. While there was descriptive evidence suggestive that the reduction in talking on any phone was more pronounced in Illinois than in control states (**Figure 1 (e)**), the corresponding DID estimate was not statistically significant at the common cutoff of 0.05.

### 4.1: Limitations

Our study, like all population-based comparison studies had several limitations. First, due to the timing of laws and sample sizes, we only analyzed a single state that enacted a comprehensive handheld phone ban. The associations we observed in Illinois may not apply to other states. Aside from underlying differences in their populations, policy effectiveness may also depend on factors such as public awareness and enforcement. Second, our analyses were unweighted. Our models adjusted for all factors associated with survey sampling and weighting, which improves our ability to infer about our target population,^27,28^ however our results should be interpreted with this caveat in mind. Third, we did not consider the potential for differential policy effects among subgroups of the population. Stratifying the analyses by, for example age, income, and education, was untenable due to the relatively small sample size in Illinois, however we view this as a promising avenue for future work. Fourth, because we selected states that neighbored Illinois, spillover effects may have undermined our ability to draw causal conclusions from our results. Because control states’ population densities are typically low near the Illinois border, we believe that spillover effects are of minimal concern in this application. An exception to this occurs in Wisconsin, in which Milwaukee is a major city located near Illinois border and Chicago, however this would encompass a small minority of the data from control states. Finally, interpreting our estimates as causal ones also requires that the parallel trends assumption holds. Although we took multiple approaches to address this assumption, we cannot formally prove that it was met. Even the parallel trends assumption was not met, the estimates in this study were nonetheless robust measures of association, which controlled for both measured and some unmeasurable confounding.

## 5: CONCLUSIONS

Our results suggest that Illinois’ comprehensive handheld phone ban reduced talking on handheld phones among drivers. Furthermore, we found evidence to suggest that the ban lead drivers who talk on the phone while driving to prefer handsfree phones over handheld ones. These findings in tandem corroborate the substitution hypothesis and suggest that Illinois’ comprehensive handheld phone ban was effective in reducing distracted driving.

## Data Availability

The study's survey data can be obtained by contacting the AAA Foundation for Traffic Safety and submitting a data request form. Programs for data processing and analysis can be obtained by contacting the corresponding author.

## 6: PRACTICAL APPLICATIONS

Our results support comprehensive handheld phone bans to reduce distracted driving, while also providing insights on how these laws associate with increasingly available handsfree technology. The practical applications of these findings are similar to those of Benedetti et al (2022): in the absence of a national prohibition of handheld phone use while driving, the findings should encourage other states to enact comprehensive handheld phone bans to improve traffic safety.

## Acknowledgements and Declarations

The authors sincerely thank the AAA Foundation for Traffic Safety, especially Dr. Woon Kim, for assistance in procuring the TSCI data and for providing valuable feedback on the manuscript.

This research did not receive any specific grant funding from funding agencies in the public, commercial, or not-for-profit sectors.

## Declarations of interest

none

The manuscript *Talking on handsfree and handheld cellphones while driving in association with handheld phone bans*, uses similar data and outcomes, with different scope and study design. It is currently under review for the Journal of Safety Research.

