## Supplemental Materials for "The impact of Illinois’ comprehensive handheld phone ban on talking on handheld and handsfree cellphones while driving"

**Table S1** presents cross tabulations of our demographic variables by exposure group and time period, with the goal of assessing covariate balance between the exposure groups both before and after the intervention. While the exposure groups are relatively similar with respect to demographics, we observed pre-to-post intervention differences in demographics within both Illinois and the control states, especially with respect to age and household size. The percentages in **Table S1** are within-sample statistics that need not reflect real-world changes in the population over time, rather they reflect changes in the samples collected for TSCI. This table underscores the importance of adjusting for demographic factors in our model, as otherwise these differences may have confounded estimates of policy associations. Second, despite the differences in demographics, there appears to be sufficient overlap between the treatment and control groups that including demographic terms in our model should suffice to account for potential confounding due to these factors.

**Table S1:** Cross-tabulations of demographic and exposure variables. “Pre-law” refers to drivers who took the TSCI survey before 1/1/2014, and “Post-law” refers to drivers who took the survey after 1/1/2014.

| Variable | Level | Illinois | | Control States | |
| --- | --- | --- | --- | --- | --- |
|  |  | Pre-law | Post-law | Pre-law | Post-law |
| Gender | Male | 48.1 | 47.4 | 50.0 | 44.9 |
|  | Female | 51.9 | 52.6 | 50.0 | 55.1 |
| Age | 21-29 | 15.9 | 9.3 | 12.9 | 8.5 |
|  | 30-44 | 21.5 | 24.8 | 21.3 | 25.1 |
|  | 45-59 | 28.8 | 46.6 | 33.0 | 40.8 |
|  | 60 or older | 33.9 | 19.4 | 32.7 | 25.7 |
| Income | <$25,000 | 12.0 | 11.9 | 18.9 | 14.8 |
|  | $25,000-$49,999 | 21.0 | 19.6 | 24.1 | 20.8 |
|  | $50,000-$74,999 | 20.2 | 20.6 | 21.6 | 21.3 |
|  | $75,000-$99,999 | 15.5 | 14.1 | 15.5 | 16.3 |
|  | $100,000 or more | 31.3 | 33.9 | 19.9 | 26.9 |
| Race/ ethnicity | Non-Hispanic White | 74.7 | 68.8 | 89.1 | 86 |
|  | Non-Hispanic Black | 14.6 | 10.9 | 4.0 | 6.0 |
|  | Non-Hispanic Other | 1.7 | 4.6 | 1.6 | 1.6 |
|  | Hispanic | 6.4 | 13.3 | 3.0 | 3.8 |
|  | Non-Hispanic 2+ races | 2.6 | 2.4 | 2.2 | 2.6 |
| Education | Less than high school | 6.0 | 6.3 | 8.8 | 6.0 |
|  | High school | 22.7 | 18.3 | 31.5 | 26.8 |
|  | Some college | 27.9 | 32.7 | 32.4 | 30.5 |
|  | Bachelor’s or higher | 43.3 | 42.7 | 27.2 | 36.7 |
| Metro. Status | Non-metro | 9.4 | 10.7 | 24.9 | 28.4 |
|  | Metro | 90.6 | 89.3 | 75.1 | 71.6 |
| Household Size | 1 | 15.0 | 11.7 | 19.3 | 12.2 |
|  | 2 | 40.8 | 26.4 | 45.8 | 32.4 |
|  | 3 | 20.2 | 18.1 | 12.2 | 19.6 |
|  | 4 | 12.9 | 22.4 | 11.8 | 19.1 |
|  | 5+ | 11.2 | 21.4 | 10.9 | 16.8 |
